# Training and reployment of non-specialists is an effective solution for the shortage of health care workers in the COVID-19 pandemic

**DOI:** 10.1101/2020.07.17.20156117

**Authors:** Ming Kuang, Jianfeng Wu, Yifeng Luo, Han Xiao, Ruiming Liang, Wenjie Hu, Shouzhen Cheng, Qian Zhou, Sui Peng, KarKeung Cheng, Haipeng Xiao

## Abstract

**Importance:** In the COVID-19 pandemic many countries encounter problems arising from shortage of specialists. Short intensive training and reployment of non-specialists is an option but the effectiveness is unknown.

**Objective:** To investigate whether there was difference in in-hospital mortality rates between COVID-19 patients managed by a mixed team (including non-specialists who had short intensive training and operated to a strict protocol) and those managed by a specialist team of health care workers.

**Design:** Cohort study, from January 26, 2020 to April 7, 2020, follow up to April 7, 2020.

**Setting:** Multicenter - Wuhan Hankou Hospital and Wuhan Xiehe Hospital, Wuhan, China.

**Participants:** 261 HCWs deployed to Wuhan from Guangdong emergency rescue team and the 269 COVID-19 patients they treated.

**Exposure:** Among 261 health care workers, 130 were in the specialist team and included 33 physicians, 32 of whom (97.0%) of whom were from relevant specialties. Each physician was in charge of 25-27 beds, with a 6-hour shift time. The mixed team included 131 health care workers, with 7 of the 28 physicians (25.0%) from relevant specialties. Each physician managed 12-13 beds, with a 4-hour shift time.

Non-specialists received short-term intensive training and then followed strict management protocols. Specialists practiced as normal.

**Main Outcomes and Measures:** Main outcome was in-hospital mortality of COVID-19 patients. Another outcome was rate of SARS-CoV-2 infection in health care workers.

**Results:** A total of 269 patients were included (144 male). In-hospital mortality rate of patients treated by the specialist teams and the mixed teams was 12.6% (20/159) and 12.7% (14/110) respectively (Difference = −0.1%, 95% CI −8.2% to 7.9%, p=.97). None of the health care workers were infected.

**Conclusions and Relevance:** Training and reployment of non-specialists is an effective solution for the shortage of health care workers in the COVID-19 pandemic.

**Key Points:** *Question:* Was there difference in mortality rates between COVID-19 patients managed by a mixed team (including non-specialists who had short intensive training and operated to a strict protocol) and those managed by a specialist team of health care workers (HCWs)?

*Findings:* In-hospital mortality rate among patients managed by specialist team (130 HCWs, 159 patients) and mixed team (131 HCWs, 110 patients) was 12.6% (20/159) and 12.7% (14/110) respectively (Difference = −0.1%, 95% CI −8.2% to 7.9%, p=.97).

*Meaning:* With shortage of specialist HCWs, training and reployment of non-specialists is an effective option in the management of COVID-19 patients.

## Background

The COVID-19 pandemic has overwhelmed medical systems worldwide. Health care workers (HCWs) from relevant specialties (eg critical care medicine and pulmonary medicine) are particularly badly affected,^1^ while those from many other departments experienced sharp declines in clinical activities^2,3^.

China was the first to face this challenge. In the first phase of COVID-19 pandemic, emergency rescue medical teams outside the epicenter of Wuhan were composed of specialists. With the shortfall in numbers, HCWs from specialties normally not directly involved in management of a condition like COVID-19 were recruited to join the rescue teams as an emergency measure. Here we describe the experience of short-term training and redeployment of a team of non-specialist HCWs sent to Wuhan from Guangzhou in January/February 2020 and the effectiveness of this initiative.

## Methods

### Participants

This study retrospectively collected information on all HCWs of two medical teams during the COVID-19 outbreak in Wuhan. One was a specialist team of the first batch of Guangdong emergency rescue team, with physicians all from COVID-19 relevant specialties, including the department of critical care medicine, pulmonary disease, emergency medicine and infectious disease. The other was the mixed team of the third batch of the First Affiliated Hospital of Sun Yat-sen University, with both specialists and non-specialists. For these HCWs, we collected information on age, gender, department, seniority, working hours and number of beds managed.

We also collected information from consecutive COVID-19 patients treated by the two teams. The inclusion criteria were: a) clinical or laboratory-confirmed COVID-19 patients^4^; b) admitted during the period the medical teams were deployed. We collected basic characteristics including age, gender, co-morbidities, laboratory tests and mortality.

This study was approved by the Ethics Committee of the First Affiliated Hospital of Sun Yat-sen University, Wuhan Xiehe Hospital and Wuhan Hankou Hospital. The informed consent was waived.

### Short-term intensive training program

HCWs from the specialist team were trained and assessed on-site for the usage of personal protection equipment (PPE) and infection control, especially during aerosol-generating procedures. Key aspects of the training, including use of PPE and ventilator, were incorporated into online videos for repeated learning (available at http://en.gzsums.net/yyxw/info_37_itemid_1235.html).

Non-specialists received a hierarchical training course including two modules: a) diagnosis procedure of COVID-19 and personal protection; b) advanced life support technology (eg. ventilator and extracorporeal membrane oxygenation (ECMO)). The videos were also made available to the mixed team. Detailed training arrangements were listed in Table 1.

**Table 1.**
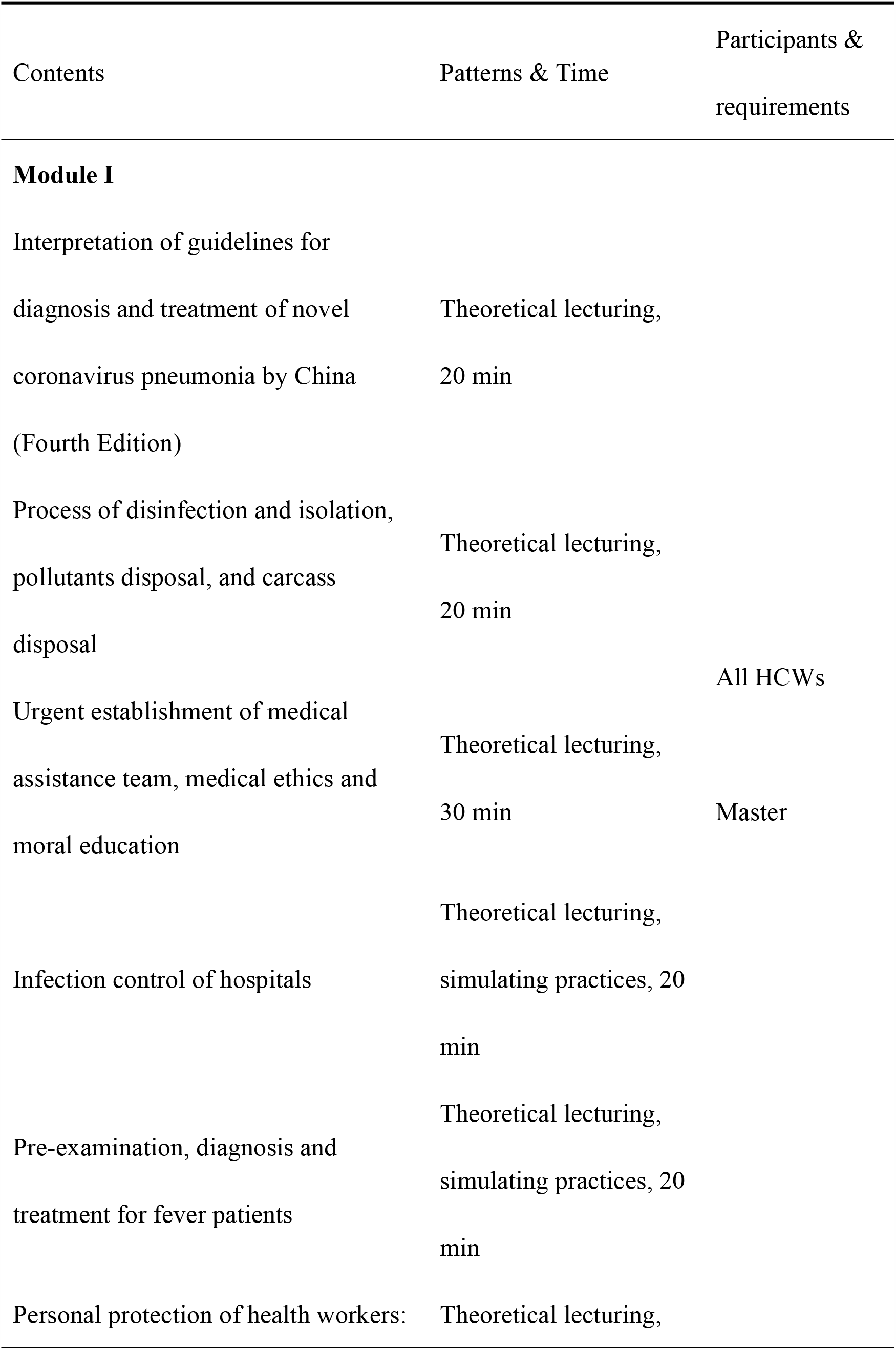
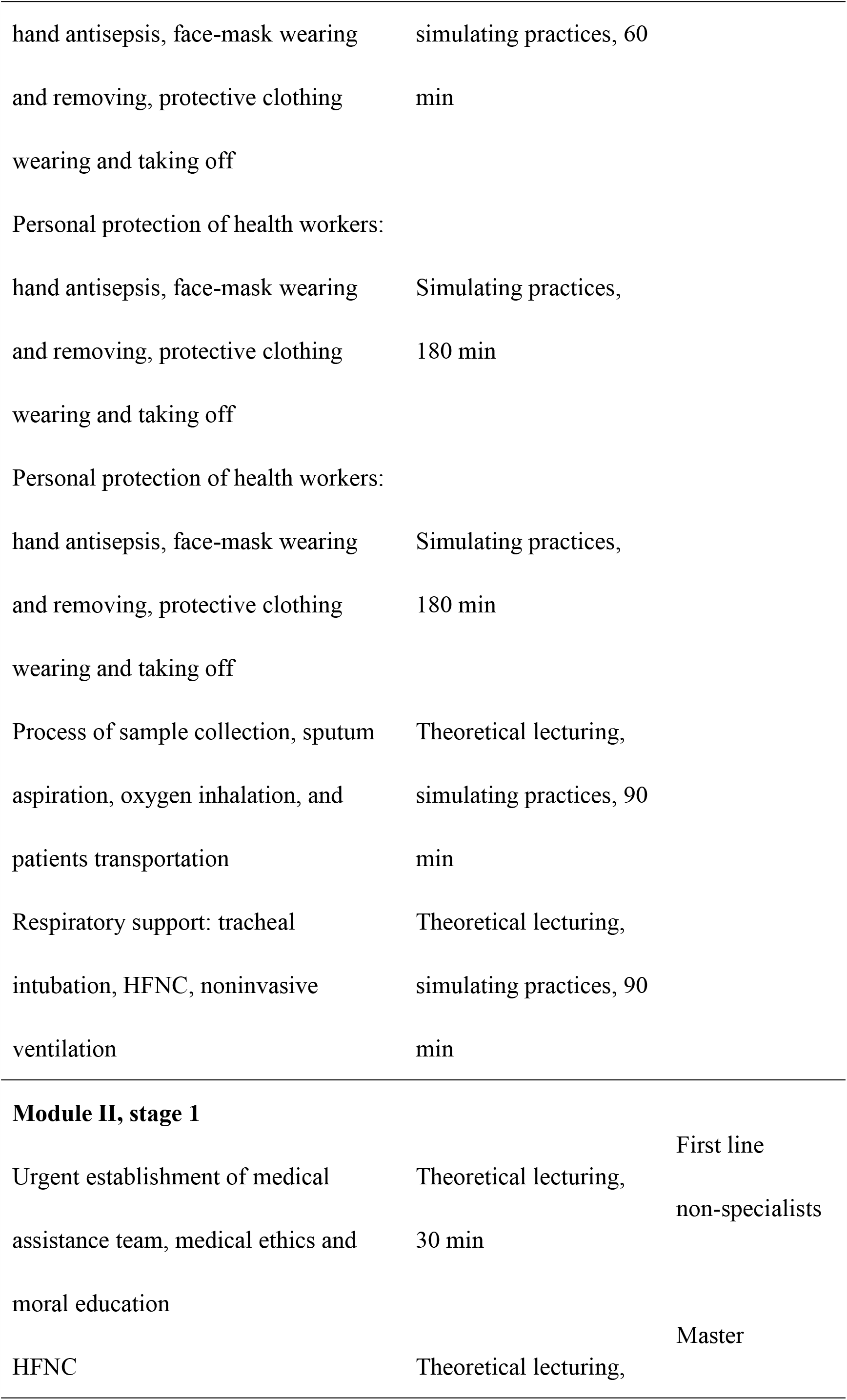
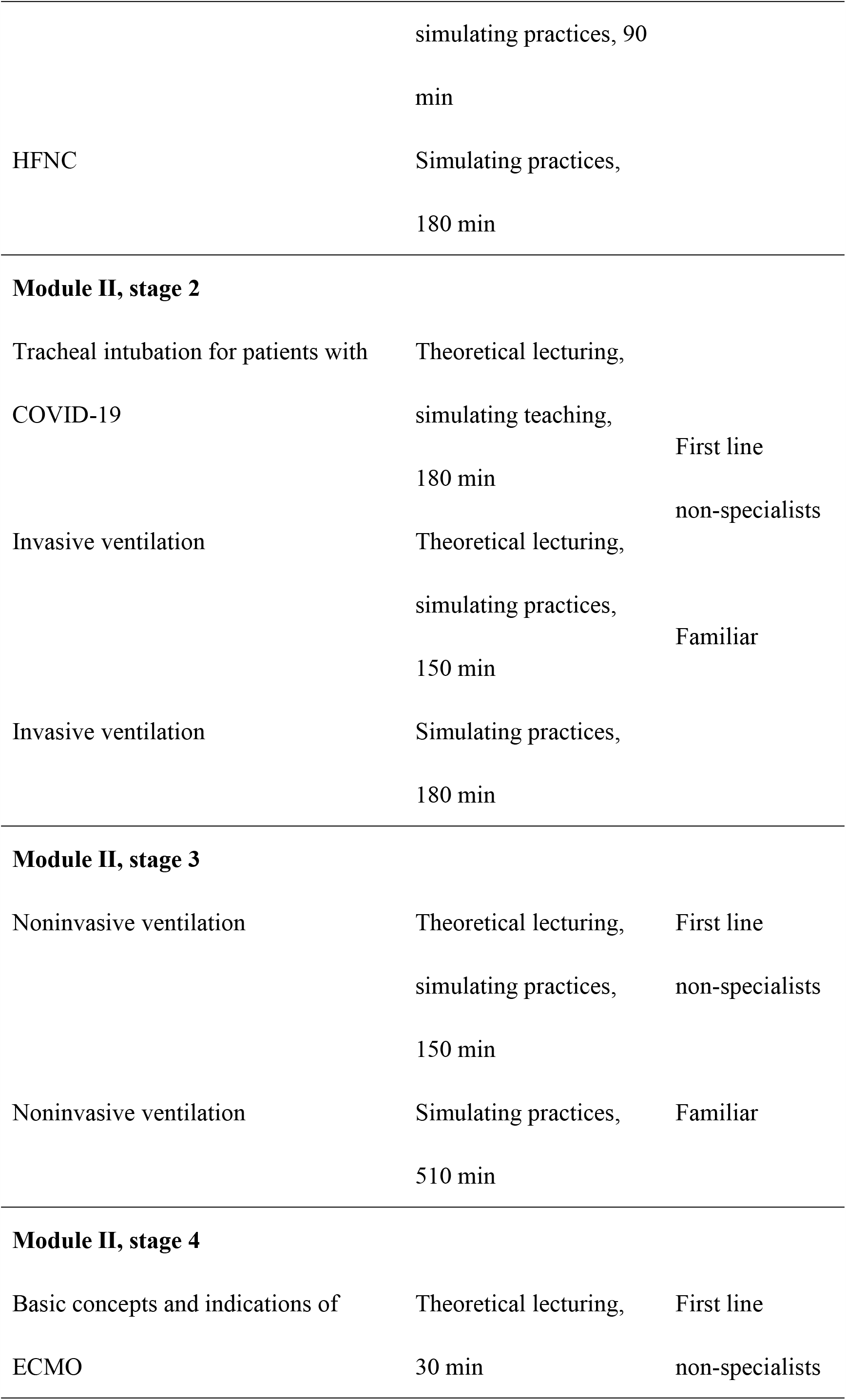
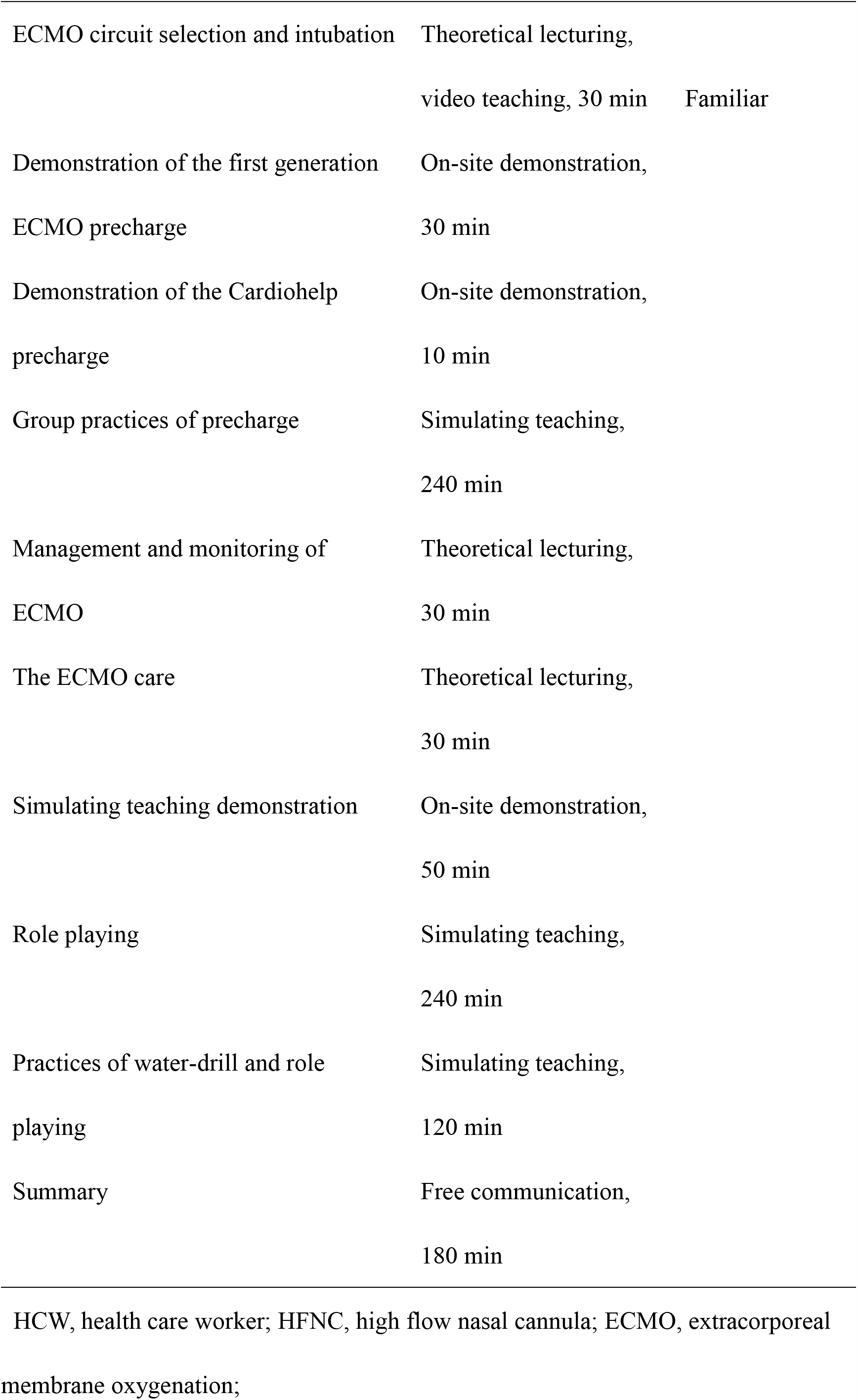
Arrangement of the short-term intensive training courses.

Each HCW of both teams was assessed by two senior specialists at the end of training and before deployment. Specialists were required to master all the contents of the two modules. Non-specialists were required to master the first module and high flow nasal cannula, and to be able to recognize indications for mechanical ventilator and ECMO.

### Medical management protocols

The specialist team was based in Wuhan Hankou Hospital and was responsible for one of the COVID-19 departments, which included 81 beds in three groups with 25-27 beds in each group. A total of 130 HCWs were in the specialist team from January 26 to March 19. There were 33 physicians, 32 (97.0%) of whom were from relevant specialties, and the remaining one isa cardiologist from the department of cardiology, with previous working experience in intensive care. Three specialists were in charge per shift. Formulation of treatment plan was led by one of the three senior specialists deployed. The mixed team operated in Wuhan Xiehe Hospital and was responsible for one department that included 50 beds in two groups with 25 beds in each group. There were 4 HCWs in each shift, with at least one specialist. The mixed team worked in Wuhan from February 8 to April 7 with a total of 131 HCWs. There were 28 physicians, and only 7 (25.0%) were from relevant specialties. The HCWs worked in pairs and managed one group of patients together. Formulation of treatment plan was led by two specialists, one of whom was among the senior specialists deployed. Should there be unexpected worsening of conditions or the need for ventilation or ECMO, specialists had to be consulted.

### Statistical analysis

Categorical variables were compared using chi-square test or Fisher exact test, and continuous variables were compared using independent sample t test. Primary outcome was in-hospital mortality of COVID-19 patients. Another outcome was rate of SARS-CoV-2 infection in health care workers. We used SAS version 9.4 for data analysis. A two-sided p-value of less than .05 was considered statistically significant.

## Results

A total of 269 patients were included, of which 159 were treated by the specialist team and 110 were treated by the mixed team. The clinical characteristics of the two groups of patients were comparable at baseline (Table 2), except that in the mixed group more patients had hypertension (50.9% vs. 38.4%, p=.04) and diabetes (32.7% vs. 20.8%, p=.03), and there were also more patients with critical condition.

**Table 2.** Baseline characteristics of patients with COVID-19 managed by the specialist and the mixed teams.

| Variable | Specialist team<br>(n=159) | Mixed team<br>(n=110) | *P*<br>value |
| --- | --- | --- | --- |
| Gender, *n* (%) |  |  |  |
| Male | 86 (54.1) | 58 (52.7) | .83 |
| Female | 73 (45.9) | 52 (47.3) |  |
| Age, median (IQR), yrs | 66 (54-72) | 63 (56-70) | .48 |
| Severity classification, *n* (%) |  |  | <.001 |
| Mild | 13 (8.2) | 7 (6.4) |  |
| Severe | 133 (83.6) | 69 (62.7) |  |
| Critical | 13 (8.2) | 34 (30.9) |  |
| Comorbidities, *n* (%) |  |  |  |
| Hypertension | 61 (38.4) | 56 (50.9) | .04 |
| Diabetes | 33 (20.8) | 36 (32.7) | .03 |
| Neoplasm | 6 (3.8) | 6 (5.5) | .56 |
| Chronic kidney disease | 7 (4.4) | 6 (5.5) | .69 |
| Nervous system disease | 10 (6.3) | 10 (9.1) | .39 |
| Respiratory system disease | 10 (6.3) | 12 (10.9) | .17 |
| Digestive system disease | 6 (3.8) | 3 (2.7) | .74 |
| Laboratory findings, median<br>(IQR) |  |  |  |
| Neutrophil count, $\times 10^9/L$ | 4.5 (2.9-7.6) | 4.9 (3.1-6.9) | .38 |
| Lymphocyte count, $\times 10^9/L$ | 0.9 (0.6-1.4) | 1.1 (0.8-1.6) | .004 |
| C-reactive protein, mg/L | 30.7 (3.8-36.9) | 15.5 (4.9-43.5) | .50 |
| Lactate dehydrogenase, U/L | 226.5<br>(178.0-345.5) | 239.5<br>(187.0-350.0) | .30 |

IQR, interquartile range.

In-hospital mortality rate of patients treated by the specialist teams and the mixed teams was 12.6% (20/159) and 12.7% (14/110) respectively (Difference = −0.1%, 95% CI −8.2% to 7.9%), p=.97).

None of the HCWs were infected in either team.^5^

## Discussion

We found that with short-term intensive training and a strict management protocol, in-hospital mortality rate among patients managed by the mixed team comprising physicians from specialties not directly related to COVID-19 was similar to that of the specialist team.

HCWs from COVID-19 relevant specialties are facing enormous physical and mental stress^6^ under the pandemic. Various policies were proposed to reploy available medical workforce ^3,7,8^, yet we could not identify any practical implementation plan in the literature. Our results suggest that brief but systematic training of non-specialists and medical management protocol for non-specialists working with a specialist could result in outcomes comparable to those of specialists. No HCW was infected and this was discussed in a previous report. ^5^.

Hierarchical training program according to the role and specialties of non-specialists is crucial. It is unrealistic to require non-specialists to master all complicated knowledge in a short time. One example of priority of learning objectives in pre-deployment training concerns ventilation and ECMO. For non-specialists the key issue is to recognize worsening of conditions and the need for ventilation or ECMO but to master details of the operation of ventilator and ECMO requires much more experience and professional knowledge. As only 2-3% of patients required ECMO use,^9,10^ the protocol required the non-specialists to consult their specialist colleagues if they came across possible indications.

As this study was not randomized, and the two medical teams did not commence their stays in Wuhan at the same time, the patients managed by the two team may have dissimilar conditions on admission. However, comparisons on some known important variables did not show major differences. Indeed there was a higher proportion of critical patients managed by the mixed team.

We conclude that training and reployment of non-specialists is an effective option in the management of COVID-19 patients in systems facing acute shortage of specialists.

## Data Availability

Haipeng Xiao had full access to all the data in the study and takes responsibility for the integrity of the data and the accuracy of the data analysis.

## Acknowledgements

### Author Contributions

M Kuang, JF Wu, YF Luo, H Xiao, RM Liang and WJ Hu contributed equally to this article. HP Xiao and S Peng supervised the study. HP Xiao, S Peng, M Kuang, JF Wu, YF Luo, H Xiao, RM Liang and KK Cheng designed the study. SZ Cheng, and WJ Hu helped to organize the study. M Kuang, JF Wu, YF Luo, WJ Hu, SZ Cheng collected the data. RM Liang and Q Zhou did the data analysis. HP Xiao, S Peng, M Kuang, JF Wu, H Xiao, WJ Hu and RM Liang wrote the draft report. HP Xiao, S Peng, KK Cheng, M Kuang, JF Wu, YF Liu, H Xiao, SZ Cheng and Q Zhou performed critical revision on the manuscript. All authors contributed to the analysis and interpretation of data. All authors approved the final version before submission.

### Funding

None

### Conflict of Interest Disclosures

We declare no conflict of interest.

### Previous Presentation of the Information Reported in the Manuscript

A manuscript describing the infection rate of the health care workers has been published in the BMJ (https://www.bmj.com/content/369/bmj.m2195).

We thank all the hospital staff members for their efforts in working in the front line.

